# Alzheimer’s Disease Classification Using 2D Convolutional Neural Networks

**DOI:** 10.1101/2021.05.24.21257554

**Authors:** Gongbo Liang, Xin Xing, Liangliang Liu, Qi Ying, Ai-Ling Lin, Nathan Jacobs

**Affiliations:** Eastern Kentucky University, Richmond, KY, USA; University of Kentucky, Lexington, KY, USA; Henan Agricultural University, Zhengzhou, Henan, China; University of Iowa, Iowa City, IA, USA

**Keywords:** CNN, 3D, MRI, Diagnosis

## Abstract

Alzheimer’s disease (AD) is a non-treatable and non-reversible disease that affects about 6% of people who are 65 and older. Brain magnetic resonance imaging (MRI) is a pseudo-3D imaging modality that is widely used for AD diagnosis. Convolutional neural networks with 3D kernels (3D CNNs) are often the default choice for deep learning based MRI analysis. However, 3D CNNs are usually computationally costly and data-hungry. Such disadvantages post a barrier of using modern deep learning techniques in the medical imaging domain, in which the number of data can be used for training is usually limited. In this work, we propose three approaches that leverage 2D CNNs on 3D MRI data. We test the proposed methods on the Alzheimer’s Disease Neuroimaging Initiative dataset across two popular 2D CNN architectures. The evaluation results show that the proposed method improves the model performance on AD diagnosis by 8.33% accuracy or 10.11% auROC, while significantly reduce the training time by over 89%. We also discuss the potential causes for performance improvement and the limitation. We believe this work can serve as a strong baseline for future researchers.

## I. Introduction

Alzheimer’s disease (AD) is a disease that affects approximately 29.8 million people worldwide in 2015 [1]. In 2018, US official death certificates recorded 122,019 deaths from AD, making AD the sixth leading cause of death in the United States and the fifth leading cause of death among Americans age 65 and older [2]. Currently, no treatment can stop or reverse the progression of AD [3]. Thus, early diagnosis is crucial for Alzheimer’s disease.

Brain magnetic resonance imaging (MRI) is the imaging modality that is widely used for AD diagnose. An MRI is a pseud-3D image composed of 2D imaging slices (Figure 1 Left). The voxels in MRIs are corresponding to the physical locations in patients’ brains. Conventional computer-aided diagnosis tools for AD classification rely on using pre-defined, hand-crafted features. However, ADs are often heterogeneous. Thus, pre-defined features may not be robust enough for modeling various AD phenotypes. Convolutional neural networks (CNN), as a promising tool, are rapidly applied in the medical imaging domain recently [4]–[11]. Compared with traditional methods, CNNs learn features directly from images, which makes CNN features more robust than pre-defined features.

**Fig. 1:**
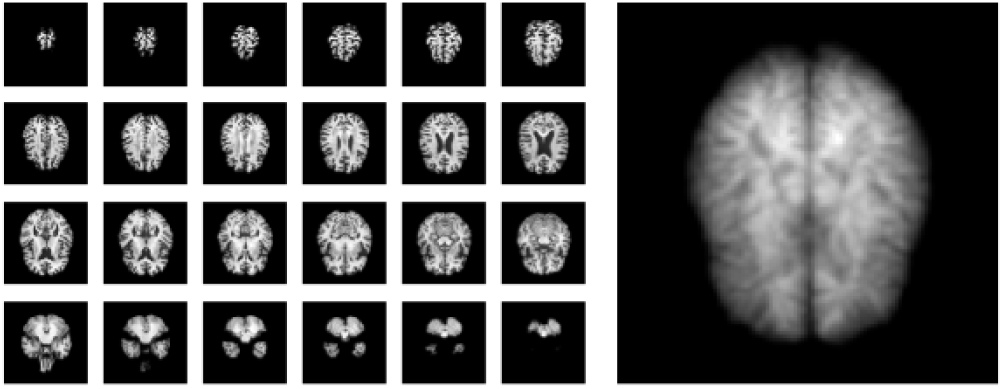
A skull-stripped brain MRI (left) and the corresponding dynamic image (right).

Korolev et al. proposed the first CNN model for AD classification in 2017 [12]. Their method uses two custom 3D CNN models for AD classification on AD classification using the ADNI dataset. The networks achieve similar performance with traditional AD classification methods that utilizing hand-crafted features. Cheng et al. [13] suggested to ensemble multiple 3D CNNs for AD classification. They, firstly, extracts local image patches from the whole image. Then, multiple 3D CNNs are trained using local patches from different locations separately. Finally, an FC layer is added on top of the multiple 3D CNNs for final prediction. Yang et al. [14] introduced an explainable version of [12] by using class activation mapping methods [15], [16]. All the existing methods are using 3D CNN networks as the building block for AD classification. However, it is well-known that 3D CNNs are computationally costly and hard to be optimized with small datasets [17], [18].

In this study, we propose to use 2D CNN models as alternative approaches for MRI classification. The methods leverage 2D CNN models on 3D imaging data by using different fusion strategies. We evaluated the proposed methods on the Alzheimer’s Disease Neuroimaging Initiative (ADNI) dataset [19] across two popular architectures. Our experimental result shows that 2D CNN models can achieve similar or better results compared with 3D CNNs, while significantly reducing the model computational cost by reducing over 89% training time. We consider our contributions to this work as the following:

- propose using 2D CNNs as an alternative approach for AD classification using 3D MRI;
- improve the AD classification performance by 8.33% accuracy or 10.11%, while reducing the training time up to 89%;
- discuss the proposed method in detail and provide a clear research direction to future researchers.

## II. Approach

The key idea of the proposed method is to convert 3D imaging data to a 2D related format using various fusion strategies. The conversion can be done at the image-level or the feature-level by using different temporal pooling methods.

### A. Temporal Pooling

Temporal pooling can be used for converting 3D MRIs to 2D images by replacing the values on the temporal dimension (or the slice dimension) with a single value. For instance, given an MRI, *I*, with the shape of *W* × *H* × *Z* (*Z* ≥ 1), a temporal pooling method, *P*, is applied to the *Z*-dimension of *I*. After the temporal pooling operation, the output shape of *P* (*I*) is *W* × *H* × 1. Temporal pooling can be applied on both the image-level and the feature-level. Two types of temporal pooling methods are used in this study: 1) max-pooling and 2) dynamic image pooling.

#### 1) Max-Pooling

Max-pooling is one of the most commonly used pooling operations using the maximum value from a region to represent the region. Figure 2 shows an demonstration of max-pooling on the 2D space. A max-pooling operation, (*P*_*max*_), is applied to an image with shape of 9× 9. The max-pooling operation has a receptive 3 × 3 kernel with stride 3 and padding 0, and the operation outputs an image with the shape of 3 × 3.

**Fig. 2:**
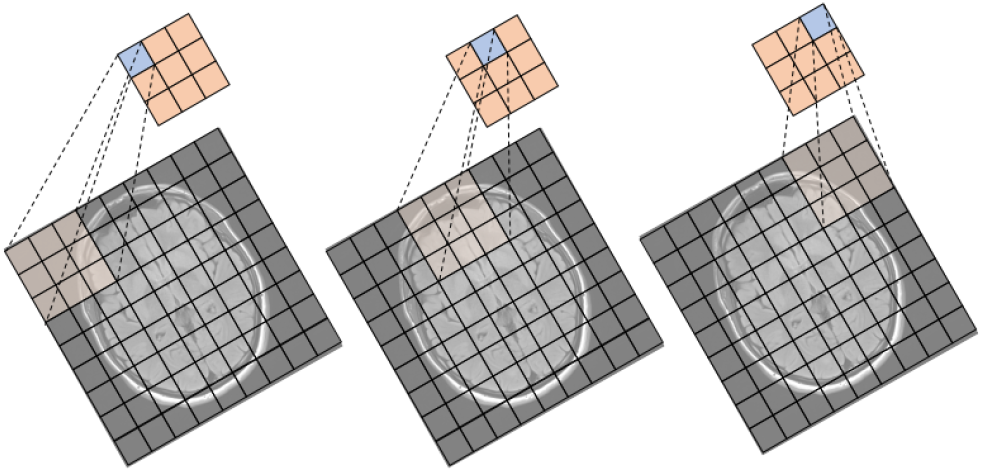
An illustration of a max-pooling process of one 3 × 3 filter (stride 3 and padding 0) from input to output.

In general, the out shape of a 2D max-pooling operation can be computed as:

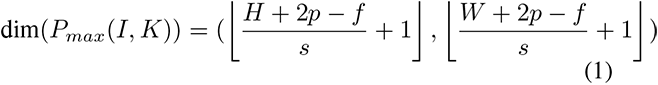

where *I* is the input image with shape of *H* × *W, K* is a 2D max-pooling function with a receptive filed of *f* × *f, p* is padding, and *s* is stride. A max-pooling operation can be applied to any dimensions. In this study, we use 1D max-pooling on the slice dimension of MRIs. The output shape of in our case is:

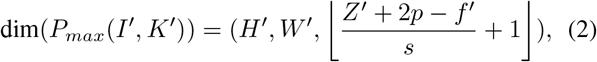

where *I*′ is an MRI with a shape of (*H*′, *W* ′, *Z*′), and *K*′ is a 1D max-pooling function with the receptive filed of *f* ′. We use *p* = 0, *f* ′ = *Z*′, and *s* = 1 in this study.

#### 2) Dynamic Image Pooling

Dynamic image pooling [20], [21] is a novel temporal pooling method that originally proposed for video clips summarization. Given a video clip, *V* = [*x*_1_, *x*_2_, *x*_3_, …, *x*_*n*_], with a shape of *w* × *h* × *n* (*n* is the number of frames). Dynamic image pooling learns a dynamic image, *µ* with a shape of *w* × *h*, that is able to rank all the frames in the video clip, such that:

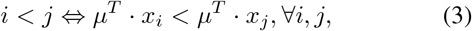

where *i* and *j* are indices of two frames. The image *µ* can be learned using RankSVM [22], [23] or any linear ranking function.

In this study, we treat MRIs as video clips and we treat each slices of an MRI as a frame in video clips. Thus, dynamic image pooling can be applied on the slice dimension of MRIs. Figure 1 Right shows an example of the output of dynamic image pooling on MRI data. Figure 3 shows an example of how to use dynamic image to rank two slices from the same MRI. More specifically, if the index of the blue slice (*Slice_B_*) smaller than the index of the green slice (*Slice_G_*), *DynamicImage* × *Slice_B_* < *DynamicImage* × *Slice_G_*.

**Fig. 3:**
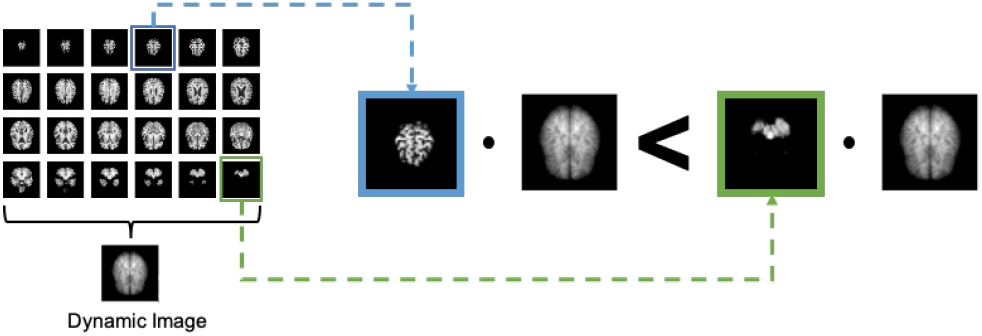
An illustration of dynamic image pooling. If the index of the blue slice (*Slice*_*B*_) smaller than the index of the green slice (*Slice*_*G*_), *DynamicImage* × *Slice*_*B*_ *< DynamicImage* × *Slice*_*G*_.

### B. Fusion Location

Usually, when utilizing 2D CNNs on 3D images, we can apply two fusion strategies that convert 3D images to 2D at two different locations: 1) early fusion and 2) late fusion. Figure 4 shows the illustrates the ideas of both strategies.

**Fig. 4:**
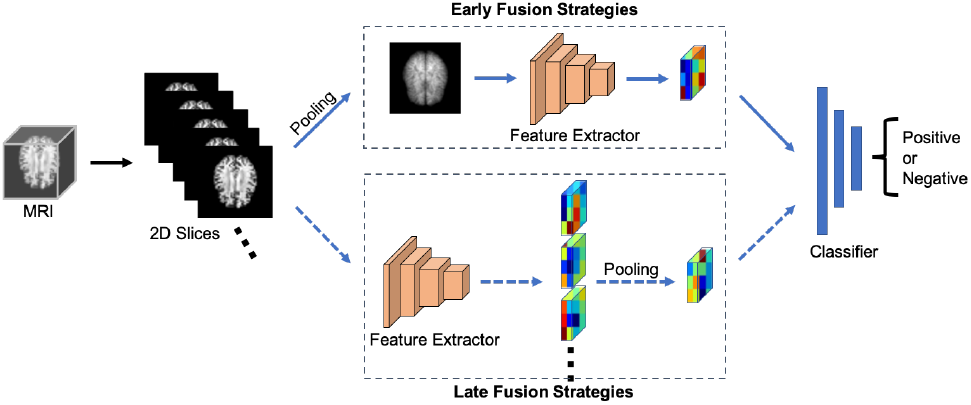
An illustration of different fusion strategies. Early fusion (Top) converts 3D images to 2D before feeding the data into a feature extractor. Late fusion (Bottom) converts 3D images to 2D after feeding the data into a feature extractor.

In early or late fusing strategies, the words “early” and “late” are respective to a CNN feature extractor. An early fusion strategy converts a 3D image to 2D before feeding the image to the feature extractor. A temporal pooling operation is usually applied on the pixel-level.

Oppositely, a late fusion strategy converts a 3D image to 2D after feeding it to a feature extractor. More specifically, each imaging slice of a 3D image is feeding into a 2D CNN feature extractor one after another. Multiple blocks of feature maps are generated at this step. Then, a temporal pooling method is applied to all the blocks of feature maps and converts them to a single block of feature maps. Finally, the fused block of feature maps is feed into the classifier for final prediction.

### C. Network Architectures and Implementation

We implement the proposed method using three different architectures with different combinations of fusion strategies and temporal pooling methods. More specifically, we have one for early fusion strategy with dynamic image pooling and two for late fusion strategies with max-pooling and dynamic image pooling, respectively.

Each architecture contains an ImageNet pre-trained CNN feature extractor and a classifier. The pre-trained feature extractor is frozen during the training stage, while the classifier is fully optimizable. The classifier contains a 1 *×* 1 Conv layer and two FC layer with 512 neurons and 2 neurons, respectively. The Conv layer aims to convert the ImageNet pre-trained features to AD-specific classification feature (Figure 5).

**Fig. 5:**
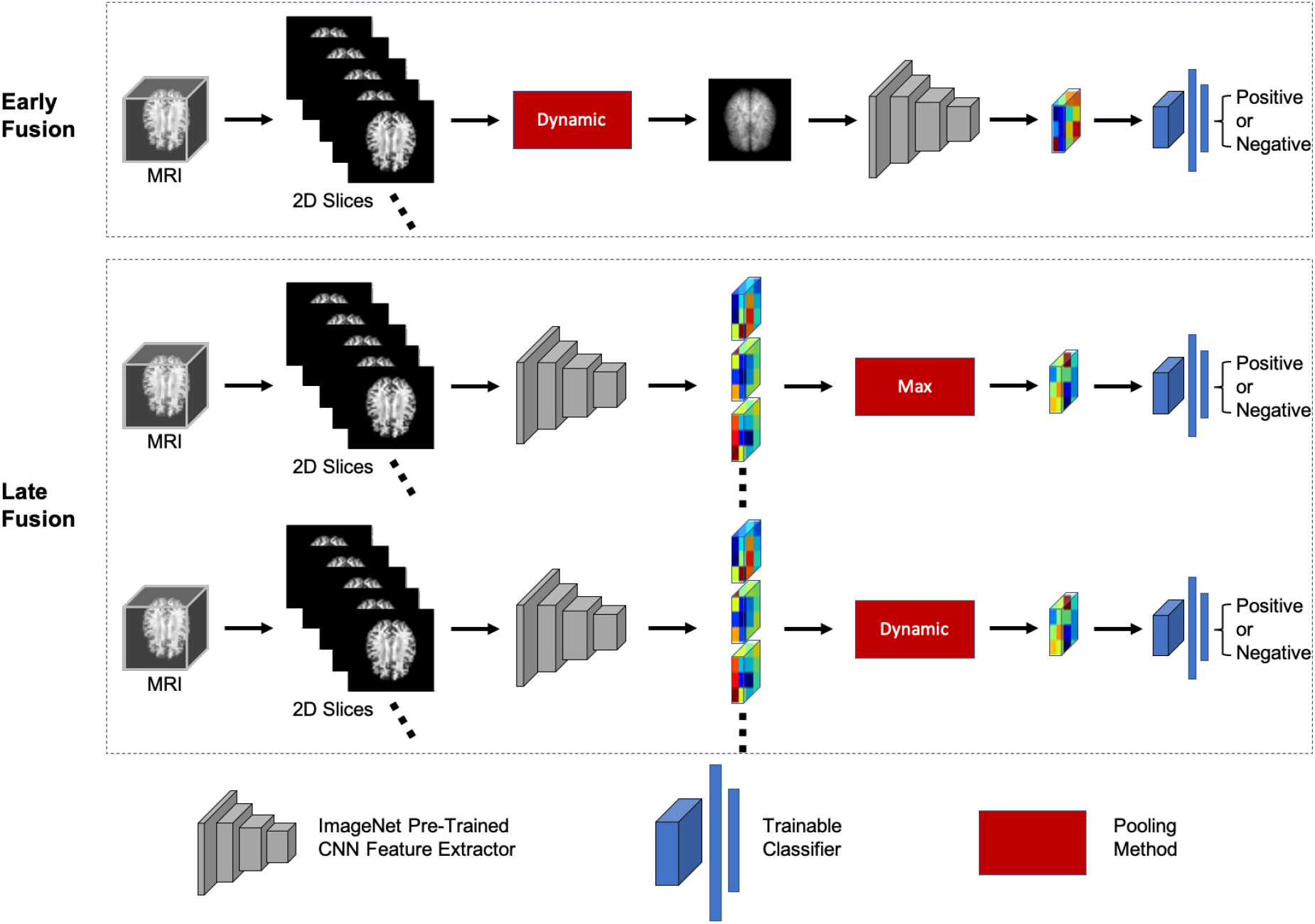
An illustration of different architectures that are used in this study.

For each architecture, we use two different backbone feature extractors, AlexNet and ResNe-18, separately. All the Conv layers of the AlexNet and ResNet-18 models are used as the feature extractors. In total, six models with different architectures are trained in this work (Table I). We implement the networks in Pytorch [24]. Weighted cross-entropy is used as the loss function. Adam [25] optimizer with learning rate of 0.0001 is used for all the models.

**TABLE I:** Detailed Architecture

| Model | Feature Extractor | Fusion Strategy | Pooling Method |
| --- | --- | --- | --- |
| **Alex_Early-Dyn** | AlexNet | Early | Dynamic Image |
| **Alex_Late-Max** | AlexNet | Late | Max-Pooling |
| **Alex_Late-Dyn** | AlexNet | Late | Dynamic Image |
| **Res_Early-Dyn** | ResNet-18 | Early | Dynamic Image |
| **Res_Late-Max** | ResNet-18 | Late | Max-Pooling |
| **Res_Late-Dyn** | ResNet-18 | Late | Dynamic Image |

## III. Evaluation

### A. Dataset

We use a subset from the ADNI dataset for our work. In total, 100 cases are used in this study, 51 cognitively normal (CN) samples and 49 AD samples. The dataset size of this study is similar to [12] and [13]. Data augmentation is applied on-the-fly for training samples, with a random combination of horizontal flip and rotations by 0, 90, 180, or 270 degrees. The data augmentation method effectively increase our training set size by a factor of 8. All the samples are skull stripped. Each sample contains a spatially normalized, masked, and N3-corrected T1 MRI with a shape of 110 × 110 × 110. We randomly split the dataset to training/testing sets with a 4:1 ratio on the patient-level. No samples for the same patient in both training and testing sets.

### B. Baseline and Evaluation Metrics

We compare our methods (i.e., the six different 2D CNN models) with [12], a 3D-ResNet model. In total, seven models are compared in this study. Each model was trained for 100 epochs with the same training/testing split. Accuracy (ACC), the area under the curve of Receiver Operating Characteristics (auROC), F1 score (F1), Precision (Prep), Recall (Recall) and Average Precision (AP) are used as the evaluation metrics.

### C. Classification Performance

Table II shows the evaluation result for all the compared models. The table reveals that four out of six our models surpass the performance of the baseline model. The best performance is achieved by **Alex** _**Late-Max**_ model, which uses AlexNet as the feature extractor and uses late fusion strategy with max-pooling. The model has a 91% accuracy and a 0.91 auROC, which is 8.33% and 10.96% higher than the baseline on accuracy and auROC, respectively.

**TABLE II:** Detailed Performance of Different Models

| Model | ACC | auROC | F1 | Prec | Recall | AP |
| --- | --- | --- | --- | --- | --- | --- |
| **3D-ResNet [12]** | 0.84 | 0.82 | 0.82 | 0.86 | 0.79 | 0.78 |
| **Alex_Early-Dyn** | 0.90 | 0.89 | 0.89 | 0.93 | 0.86 | 0.86 |
| **Alex_Late-Max** | **0.91** | **0.91** | **0.91** | **0.97** | **0.85** | **0.90** |
| **Alex_Late-Dyn** | 0.90 | 0.88 | 0.90 | 0.94 | 0.88 | 0.88 |
| **Res_Early-Dyn** | 0.83 | 0.81 | 0.82 | 0.85 | 0.80 | 0.78 |
| **Res_Late-Max** | 0.84 | 0.76 | 0.84 | 0.85 | 0.80 | 0.78 |
| **Res_Late-Dyn** | **0.88** | **0.86** | **0.86** | **0.91** | **0.85** | **0.84** |

Regarding the fusion strategies and pooling methods, there is no clear winner. However, we think late fusion with dynamic image pooling is generally a good combination regardless of the choice of a feature extractor. The performance of the late fusion with a dynamic image pooling method is relatively consistent among the two feature extractors, with only a 2% difference for most of the evaluation metrics. However, the performance differences between feature extractors for other fusion methods are much larger when compared between different feature extractors.

### D. Model Training Time

We train all the models using an Nvidia GTX 1080 GPU card. Each model was trained for 100 epochs. We use batch size 16 for all of our models and batch size 8 for the baseline model, which is the largest batch size we can fit into the GPU memory. Table III shows the end-to-end training time for the baseline model and ours late fusion with dynamic image pooling models.

**TABLE III:** Training Time of Different Models

| Model | Training Time (Mins) |
| --- | --- |
| **3D-ResNet [12]** | 3916 |
| **Alex_Late-Dyn** | 213 |
| **Res_Late-Dyn** | 421 |

The table reveals that our models significantly reduces the training time compared with the baseline model. The baseline model was trained for over 65 hours and achieved an 84% accuracy and a 0.82 auROC. Our model with 2D ResNet-18 backbone only needed about 7 hours and got an even better performance, such that an 88% accuracy and a 0.86 auROC.

One thing worth noting is that since we use the pre-trained feature extract fixed during our training, we can further reduce the training time by pre-generating the image feature maps. In such a way, we only need to train the classifier. According to our experiments, the classification training with pre-generated feature maps can be done within 30 minutes.

## IV. Discussion

Compared with the traditional 3D CNN approach, the proposed 2D CNN models can achieve better performance while significantly reduce the training time. We believe both the performance improvement and the training time reduction are primarily caused by reducing model complexity. Empirically, 2D CNNs usually have less trainable parameters than 3D CNN models. Thus, a 2D CNN model may require less training data and easier to be optimized. Besides, transfer learning can be easily applied to 2D CNN models since there are many large datasets are available for pre-training. However, it is difficult to apply transfer learning on to 3D CNNs due to the lack of pre-training dataset.

It is surprising that all of our models with AlexNet feature extractor are outperformed the baseline model, while only one ResNet backbone model has a better performance than the baseline. One reasonable explanation is that AlexNet is shallower than ResNet-18, with 5 Conv layers vs. 18 Conv layers, respectively. The image features extracted by an AlexNet may contain more low-level information than ResNet-18, while the features of ResNet-18 may contain more high-level information that towards the object level. Since the feature extractors are pre-trained using the natural imaging dataset and frozen during our training, the low-level information may be more informative for our project because the differences between MRIs and natural images. Thus, models using AlexNet backbone have better performances than the ones using ResNet-18 backbone.

One limitation of this work is the dataset used in this study is small. Though the size is similar with the one used in [12], the small dataset size may limit the 3D model’s performance since it may not be sufficient to tune the 3D model end-to-end. Thus, one of the future research directions of this work is to work on a large dataset.

During the experiments, we found that both of the proposed methods and the baseline method would get performance decrease when using MRIs without skull-stripping as input. One possible explanation is that the pixel values of skulls are remarkably higher than brain tissues in MRIs. During the fusion stage, it is likely that more information from skull areas is selected. However, such information may not be useful for Alzheimer’s disease diagnosis. Similarly, the feature extraction part of a 3D CNN model can also be considered as a special form of temporal pooling. Hence, the 3D method may also suffer from the same reason. Thus, another future research direction is to improve classification performance using MRIs without skull-stripping.

## V. Conclusion

In this study, we propose to use 2D CNN models combined with different temporal pooling strategies for the Alzheimer’s disease diagnosis. Compared with the conventional 3D CNN approach, the proposed method is able to improve the classification performance by 8.33% or 10.11% for the accuracy or auROC, respectively. In addition, the proposed methods reduce the training time up to 89%, from 65 hours to 7 hours. We believe the proposed methods can serve as a strong baseline for future researchers.

## Data Availability

The data used in this study is from The Alzheimer's Disease Neuroimaging Initiative (ADNI) (http://adni.loni.usc.edu)

